# Risk Factors for Developing Diabetic Foot Ulcer with Nephropathy, Diabetic Kidney Disease and Renal Failure Statistical Analysis of 10,680 Patients’ Cohort

**DOI:** 10.1101/2020.06.11.20128488

**Authors:** Kamran Mahmood Ahmed Aziz

**Affiliations:** Consultant Diabetologist; Research Scientist and Clinical Investigator Aseer Diabetes Center of Aseer Central Hospital, Ministry of Health, Abha, Saudi Arabia

## Abstract

It has been cited in the research that patients with diabetic kidney disease (DKD) and foot ulcer have poor prognosis; and foot ulceration is associated with kidney dysfunction. However, no study have found association of diabetic foot ulcer (DFU) with diabetic kidney dysfunction and their co-existing risk factors including blood pressure, serum creatinine, microalbumin, spot urine protein, HbA1c and associations of nephropathy and DKD with low ankle brachial index (ABI). We monitored, collected and analyzed the data for 10,680 patients for a duration of more than 13 years. 12% of patients demonstrated DFU. Nephropathy was observed in 39% of patients; 43% was hypertensive while 15% demonstrated DKD or CKD. Levels of HbA1c, creatinine,, systolic and diastolic blood pressures, microalbuminuria, spot urine protein, and spot urine protein to creatinine ratio were higher among the groups with foot ulcers (p-value < 0.0001 for all). Average ABI was observed to be lower among the groups demonstrating nephropathy and DKD with significant p-values (p=0.025 and 0.022 respectively). Pearson’s χ^2^ and logistic regression with odds ratio were also analyzed for DFU with HTN, nephropathy and DKD. DFU was significantly associated with HTN (odds ratio 2.2 ; 95% CI 1.66 to 2.9; p < 0.0001), nephropathy (odds ratio 4.77 ;95% CI 3.53 to 6.5; p < 0.0001) and DKD (odds ratio 4.77 and 6.83; 95% CI 4.6 to 10.2; p < 0.0001). ROC was used to find out cutoff values, sensitivity and specificity. HbA1c of 7.8% was 60% sensitive and 52% specific for the development of DFU (AUC = 0.58; 95% CI 0.521 to 0.624; p < 0.0006). Creatinine of 1.2 mg/dl was 75% sensitive and 48% specific for DFU (AUC = 0.58; 95% CI 0.640 to 0.715; p < 0.0001). Spot urine protein excretion from nephrons of 35 mg/dl was 88% sensitive and 90% specific for the development of DFU (AUC = 0.585; 95% CI 0.555 to 0.616; p < 0.0001). Our data has demonstrated the first time such associations and confirmed that nephropathy or renal failure are risk factors for the development of DFU. HbA1c should be optimal and near to the targets to improve wound healing. Our study has prompted diabetologists for regular and routine assessment of the feet and early screening of diabetic patients for neuropathy, nephropathy, hypertension, dyslipidemia and other diabetic complications as well.

## INTRODUCTION

Diabetes mellitus is a global health problem. Diabetic foot ulcer (DFU) or diabetic foot infection (DFI) are the major cause of lower extremity amputations (LEA). More than 25% of diabetic patients suffer from foot amputations during their lifetime and more than 85% of lower extremity amputation are due to foot infections or ulcerations. Furthermore, diabetes is now the most common cause of preventable Charcot neuroarthropathy. Diabetic foot problems are considered a complex group of pathologies and also known as “diabetic foot syndrome„ (DFS), including both neuropathy and vasculopahty or vascular insufficiency [1-12]. Periodic neurological examination is essential with assessment of ankle brachial index (ABI) and for foot pulses.

Diabetes is major cause of other microvascular diseases, and leading to microvascular complications such as retinopathy, nephropathy, and chronic renal failure or end stage renal disease (ESRD). Landmark diabetes control and complication trial (DCCT) and UK Prospective Diabetes Study (UKPDS) have shown that reductions in HbA1c levels will reduce the risk of diabetic macrovascular and microvascular complications [13,14].

Diabetes mellitus is a strong risk factor for chronic kidney disease (CKD). Chronic involvement of the kidney in diabetic state is currently termed as diabetic kidney disease (DKD). Initially, under the influence of high blood pressure (HTN), diabetic kidney disease results in microalbuminuria or gross proteinuria, nephropathy and may to end stage renal disease (ESRD) if diabetes is uncontrolled.

Furthermore, hypertension (HTN) or elevated systolic and diastolic pressures are also major risk factors for the nephropathy, proteinuria and ESRD. HTN and Atherosclerotic cardiovascular disease (ASCVD) are also leading cause of morbidity and mortality among diabetics, coexisting with DKD [15-24].

Moreover, research trials have demonstrated that a low ankle brachial index (ABI) of ≤ 0.9 is a predictor and risk for the coronary artery disease (CAD) and peripheral vascular disease (PVD). Research has demonstrated that Chronic renal dysfunction is also associated with low ABI and with high mortality among patients on hemodialysis [25-31].

Microalbuminuria (30-300 mg/L albumin excretion in urine) is considered a risk biomarker for the cardiovascular disease. However, it does not represent actual underlying renal injury [32-38]. Recently, spot urine protein UPr and its ratio with spot urine creatinine UCr is also recommended for diagnosing and monitoring proteinuria; this inexpensive, easily and quickly performed at tertiary care endocrine, diabetes and hospital settings. Spot urine protein estimation is very helpful when microalbumin exceeds 300 mg albumin per day (gross proteinuria). The spot urine protein/creatinine ratio (UPr/UCr or PCR) correlates well with total protein excretion per day, and provides good estimation of protein excretion from the kidney [39-49].

There are some research trials which have demonstrated association between diabetic foot ulceration and development of kidney dysfunction or failure. In the past decade, much work was done to find association between these two pathologies to prevent health cost burden [50]. However, much less work has been published on the risk of developing renal failure (CKD/DKD), associated co-morbidities, risk factors and proteinuria with DFU. There is a need to study associations between these risk factors, and development of DKD with DFU. Hence according to the literature review, we hypothesized that the risk of developing DFU with nephropathy and DKD was associated with other co-morbidities also, including HTN or increased systolic/diastolic blood pressures, increased levels of serum creatinine, microalbumin, and spot urine protein, which was not studied previously in such a detailed manner. For the assessment of glycemic control, HbA1c was also tested. Levels of these variables were measured with or without DFU and associations with HTN, nephropathy and DKD.

## STUDY DESIGN AND METHODS

This is a prospective cross sectional cohort study, conducted at the diabetology clinic of Aseer Endocrine and Diabetes Center of Aseer Central Hospital, Ministry of Health, Saudi Arabia. Study started in August 2005, until September 2018 for more than thirteen years. 10,370 diabetic patients were selected for the study. We included both type-1 and type-2 diabetic patients. Children of less than 13 years of age, patients with severe liver disease, urinary tract infection, known cases of nephrotic syndrome before the onset of diabetes, with end stage renal disease (ESRD) or dialysis and pregnant subjects were excluded from this study. Blood pressure (BP) was measured by standardized methodology. BP of ≥ 140/90 was labeled as hypertension (HTN). Patients with active foot ulcer (of any grade or severity) and regular follow up in diabetic foot clinic were labeled as DFU or DIF.

FDA approved arterial doppler ultrasonic device (atys M**è**dical Doppler System Inc. USA) was used to measure ABI. Measurements were carried in resting and supine position. Brachial pressure in right arm was measured by doppler probe (8 MHz). This was then applied to right foot arteries (dorsalis pedis or posterior tibial artery). Artery with higher pressure was recorded. Right ABI was calculated as ABI = brachial pressure / foot pressure. Same clinical method was applied to measure left ABI. Average ABI for both feet was calculated for statistical analysis with nephropathy and DKD.

## LABORATORY METHODS

Blood samples for clinical chemistry was collected in fasting state. Serum creatinine (mg/dl) was quantitatively measured by CREA methodology by Dimension® clinical chemistry device (Siemens Healthcare Diagnostics Inc. Newark, DE 19714, USA). This technique involves picrate for the measurement of creatinine in plasma and urine. In the presence of a strong base NaOH, picrate chemically reacts with creatinine to form a red chromophore. The rate of increasing absorbance at 510 nm due to the formation of this chromophore is directly proportional to the creatinine concentration in the sample of blood or urine and which, is measured using by a bichromatic (510,600nm) rate methodology. [51 52 53] Patients with serum creatinine ≥ 1.5 mg/dl were considered as CKD or DKD HbA1c was measured by A1c Flex® Reagent by the Dimension® clinical chemistry system (Siemens healthcare diagnostics Inc. Newark, DE 19714, USA). This detects *in vitro* quantitatively both percent hemoglobin A1c and total hemoglobin. The techniques is based on a turbidimetric inhibition immunoassay (TINIA) principle, and the measurement of total hemoglobin is based on a modification of the alkaline hematin reaction, an NGSP certified methodology. % HbA1c in percent was calculated by the percentage of total hemoglobin that is glycated (in g/dL), which was then standardized according to the DCCT results.

Nephropathy detection was carried out by the measurement of albumin or protein in urine. QuikCheck™ urinalysis reagent strips (ACON biotech, Co., Ltd.) was used to detect gross proteinuria (macroalbuminuria). Simply, this technique involves the phenomenon of pH indicators, releasing hydrogen ions to the protein. Samples demonstrating gross proteinuria (macroalbuminuria) by the color change of the reagent strips (from 1+ to 4+ proteins) were considered “nephropathy„. Microalbumin was detected in urine by MALB method (Dimension® clinical chemistry system device, Siemens Healthcare Diagnostics Inc. Newark, DE 19714, USA). This measures albumin *in vitro* quantitatively (mg/L) by particle-enhanced turbidimetric inhibition immunoassay (PETINIA) methodology by color change. Samples positive for microalbuminuria were labeled as nephropathy. Spot urine protein UCFP (Urinary/ Cerebrospinal Fluid Protein) was measured by Dimension® clinical chemistry system (Siemens healthcare diagnostics Inc. Newark, DE 19714, U.S.A). This detects *in vitro* total protein in human urine and cerebrospinal fluid directly and quantitatively by pyrogallol red molybdenum method (Y. Fujita, I. Mori and S. Kitano methodology) [54]. Chemically, pyrogallol red combined with sodium molybdate to form a red complex with maximum absorbance at 470 nm. The protein in the sample reacted with this complex in acid solution to form a bluish-purple colored complex, which absorbs at 600 nm. The absorbance at 600 nm is directly proportional to the concentration of protein in the sample. The analyte concentration is determined by calculation of a logit curve fit on a previously stored calibration curve. PCR (protein to creatinine ratio) was measured by the formula, PCR = spot urine protein / spot urine creatinine. Laboratory samples were reviewed by Natcom Hospital Information System (NATCOM HIS; National Computer System Co. Ltd [55].

## STATISTICAL METHODS

Clinical data was analyzed by IBM® SPSS® statistics, version 20 (IBM Corp.). Data was summarized as percentages with mean ± SD and 95% CI. Independent t-test was used to test the significance between the group of variables. Pearson chi-square (χ^2^) was used to find significant associations among DFU with HTN, nephropathy and DKD.

Logistic Regression, Odds Ratio were considered to measure associations of DFU with HTN, nephropathy, and DKD/CKD. ROC was used to find cutoff values, sensitivity and specificity for HbA1c, creatinine and spot urine protein. Statistical power of 90% and p-values (two-sided) of less than 0.05 were considered significant.

## PATIENT CONSENT

The study was reviewed and approved by the research committee of Aseer Diabetes and Endocrine Center, and all methodologies on subjects reported in were in accordance with the Helsinki Declaration of 1975 (revised in 2008).

## RESULTS

Table-1 presents demographic data. 12% of patients demonstrated diabetic foot infection. Nephropathy was observed in 39% of patients; 43% was hypertensive and15% demonstrated DKD/CKD. Descriptive statistics for variables are shown in table-2.

**Table-1.**
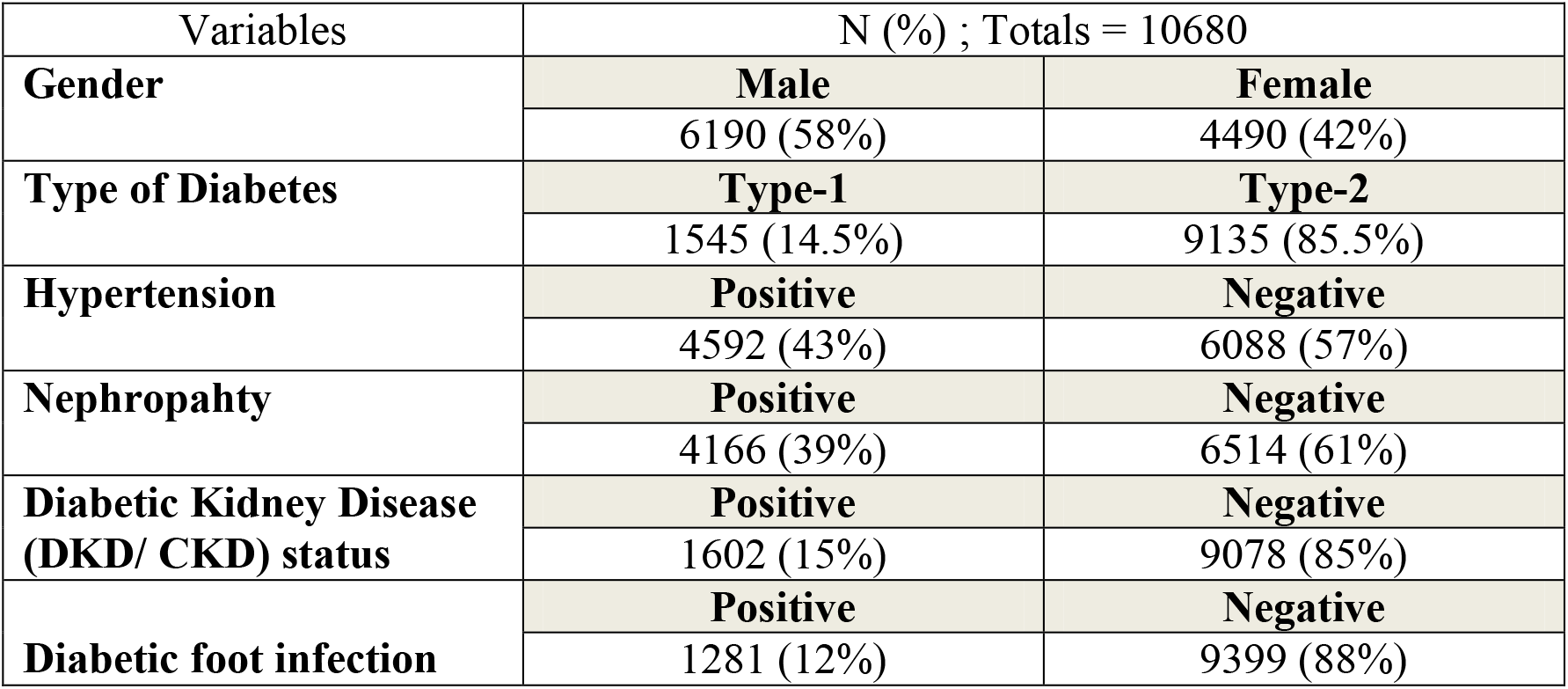
Demographic data of diabetic patients

| Variables | N (%) ; Totals = 10680 |  |
| --- | --- | --- |
| **Gender** | **Male** | **Female** |
|  | 6190 (58%) | 4490 (42%) |
| **Type of Diabetes** | **Type-1** | **Type-2** |
|  | 1545 (14.5%) | 9135 (85.5%) |
| **Hypertension** | **Positive** | **Negative** |
|  | 4592 (43%) | 6088 (57%) |
| **Nephropathy** | **Positive** | **Negative** |
|  | 4166 (39%) | 6514 (61%) |
| **Diabetic Kidney Disease (DKD/ CKD) status** | **Positive** | **Negative** |
|  | 1602 (15%) | 9078 (85%) |
| **Diabetic foot infection** | **Positive** | **Negative** |
|  | 1281 (12%) | 9399 (88%) |

**Table-2.** Variables with mean ± SD

| Variables | Mean ± SD |
| --- | --- |
| Age (years) | 54 ± 13.7 |
| Diabetes duration (years) | 16 ± 8.7 |
| Serum creatinine (mg/dl) | 0.965 ± 0.679 |
| HbA1c % (g/dl) | 7.8 ± 1.5 |
| Systolic BP (mmHg) | 129 ± 15.8 |
| Diastolic BP (mmHg) | 79.6 ± 9.3 |
| Microalbumin in urine (mg/L) | 74.6 ± 106.7 |
| Spot Urine protein (mg/dl) | 53.6 ± 29.5 |
| Spot Urine creatinine mg/dl | 121 ± 73.7 |
| Protein to creatinine ratio (PCR) | 0.61 ± 2.03 |
| Average ABI (right and left side of body) | 1.23 ± 0.33 |

Significant t-test among group of variables (HbA1c, systolic and diastolic blood pressures, microalbuminuria, spot urine protein, creatinine, and their ratio) is presented in tble-3. Levels of these variables were high among the groups demonstrating diabetic foot infection, with significant p-values.

Table-4 demonstrates average ABI values with nephropathy and DKD. Average ABI values were lower among the groups with nephropathy and DKD.

**Table-3.** T-test between groups of variables (with and without foot ulcer) with mean±SD and p-values.

| Variables and indicators | Comparison of variables with or without Foot Ulcer |  |  |  |  |
| --- | --- | --- | --- | --- | --- |
|  | Mean ± 95 % CI |  | F-value | T-value | P-values |
| **HbA1c % (g/dl)** | **With Foot Ulcer** | **Without Foot Ulcer** | 2.1 | 3.1 | 0.002 |
|  | 8.6 ± 1.61<br>8 to 8.7 | 7.8 ± 1.47<br>7.7 to 7.9 |  |  |  |
| **Creatinine (mg/L)** | **With Foot Ulcer** | **Without Foot Ulcer** | 83 | 8.85 | < 0.0001 |
|  | 1.3 ± 1.02<br>1.17 to 1.44 | 0.898 ± 0.585<br>0.87 to 0.93 |  |  |  |
| **Systolic BP (mmHg)** | **With Foot Ulcer** | **Without Foot Ulcer** | 5.5 | 8.3 | < 0.0001 |
|  | 136.5 ± 18<br>135 to 139 | 127 ± 15<br>126 to 128 |  |  |  |
| **Diastolic BP (mmHg)** | **With Foot Ulcer** | **Without Foot Ulcer** | 5.8 | 5.97 | < 0.0001 |
|  | 83 ± 10<br>82 to 95 | 78 ± 8<br>78 to 80 |  |  |  |
| **Microalbumin in urine (mg/L)** | **With Foot Ulcer** | **Without Foot Ulcer** | 99.5 | 10.5 | < 0.0001 |
|  | 170.3 ± 198<br>131.2 to 297 | 55.2 ± 78<br>49.2 to 81.3 |  |  |  |
| **Spot Urine protein (mg/dl)** | **With Foot Ulcer** | **Without Foot Ulcer** | 202 | 13.6 | < 0.0001 |
|  | 270 ± 176<br>135.6 to 440 | 39.8 ± 29<br>17.3 to 65 |  |  |  |
| **Spot Urine creatinine mg/dl** | **With Foot Ulcer** | **Without Foot Ulcer** | 0.335 | 0.189 | 0.850 |
|  | 119 ± 66<br>114.5 to 124.3 | 120.5 ± 90<br>108 to 133 |  |  |  |
| **Protein to creatinine ratio (PCR)** | **With Foot Ulcer** | **Without Foot Ulcer** | 166.8 | 11 | < 0.0001 |
|  | 1.93 ± 3.7<br>1.4 to 2.43 | 0.23 ± 0.95<br>0.16 to 0.3 |  |  |  |

**Table-4.** Significant associations between ABI and groups of variables (nephropathy and DKD) with mean±SD and p-values

| Variables and indicators | Comparison of variables with or without nephropathy and DKD |  |  |  |  |
| --- | --- | --- | --- | --- | --- |
|  | Mean ± 95 % CI |  | F-value | T-value | P-values |
| **ABI (average)** | **With Nephropahty** | **Without Nephropahty** | 5 | 1.99 | 0.025 |
|  | 1.2 ± 0.28<br>1.16 to 1.24 | 1.29 ± 0.38<br>1.22 to 1.34 |  |  |  |
| **ABI (average)** | **With DKD** | **Without DKD** | 2 | 1.7 | 0.022 |
|  | 1.15 ± 0.23<br>1 to 1.2 | 1.25 ± 0.33<br>1.2 to 1.3 |  |  |  |

Pearson’s (χ^2^) and logistic regression with odds ratio is presented in table-5. DFU was significantly associated with HTN (odds ratio 2.2 ; 95% CI 1.66 to 2.9; p < 0.0001). Similarly, DFU was significantly associated with the development of nephropathy and DKD/CKD; odds ratio 4.77 (95% CI 3.53 to 6.5; p < 0.0001) and 6.83 (95% CI 4.6 to 10.2; p < 0.0001), respectively.

**Table-5.** Significant Pearson’s (χ^2^) results for the variables HTN, nephropathy, and CKD/DKD

| **Variables** | **Pearson's ($\chi^2$) ;**<br>**p-value** | **Fisher's exact**<br>**test**<br>**p-value** | **Linear-by-**<br>**linear**<br>**Association**<br>**p-value** | **Logistic**<br>**Regression and**<br>**Odds Ratio**<br>**(95% CI)** |
| --- | --- | --- | --- | --- |
| **Diabetic foot**<br>**ulcer and HTN** | < 0.0001 | < 0.0001 | < 0.0001 | 2.2 (1.66 to 2.9) |
| **Diabetic foot**<br>**ulcer and**<br>**nephropathy** | < 0.0001 | < 0.0001 | < 0.0001 | 4.77 (3.53 to 6.5) |
| **Diabetic foot**<br>**ulcer and**<br>**DKD/CKD** | < 0.0001 | < 0.0001 | < 0.0001 | 6.83 (4.6 to 10.2) |

Table-4 (and figure 1,2, and 3) demonstrates ROC for DFU with HbA1c, creatinine and spot urine protein. HbA1c of 7.8% was 60% sensitive and 52% specific for the development of DFU (AUC = 0.58; 95% CI 0.521 to 0.624; p < 0.0006). Creatinine of 1.2 mg/dl was 75% sensitive and 48% specific for DFU (AUC = 0.58; 95% CI 0.640 to 0.715; p < 0.0001). Spot urine protein excretion from nephrons of 35 mg/dl was 88% sensitive and 90% specific for the development of DFU (AUC = 0.585; 95% CI 0.555 to 0.616; p < 0.0001).

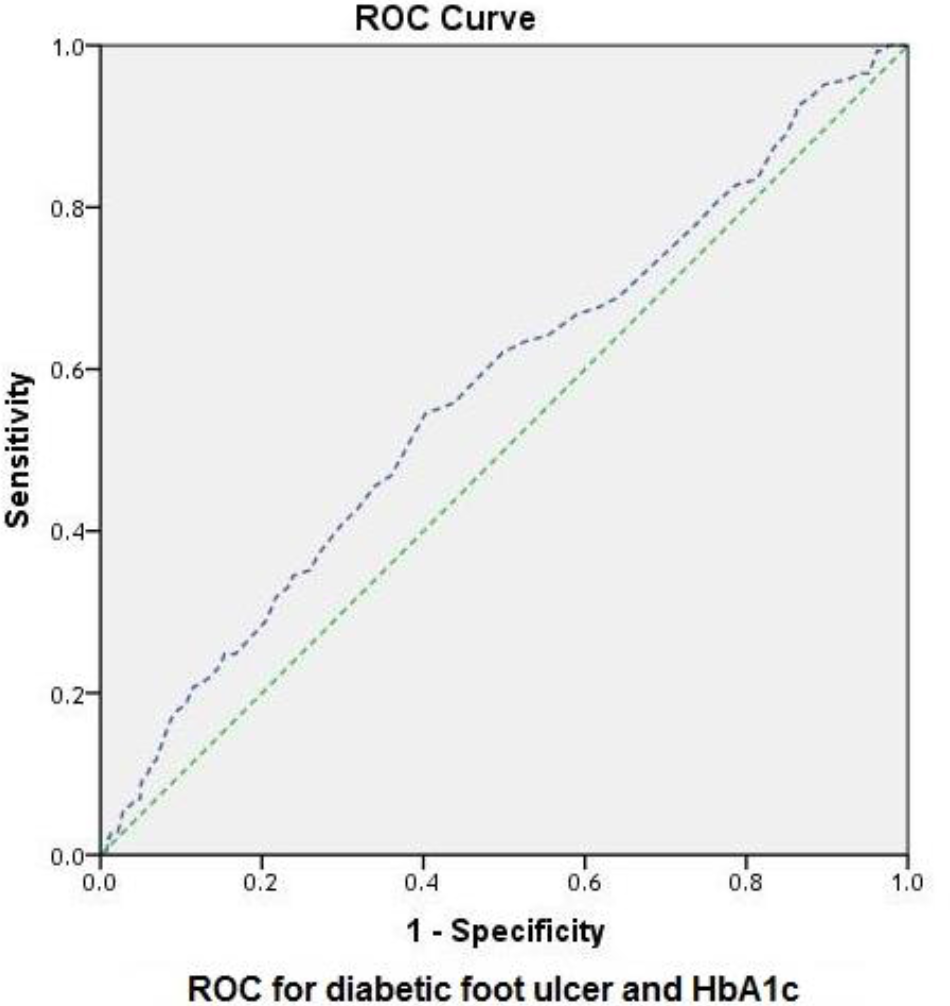

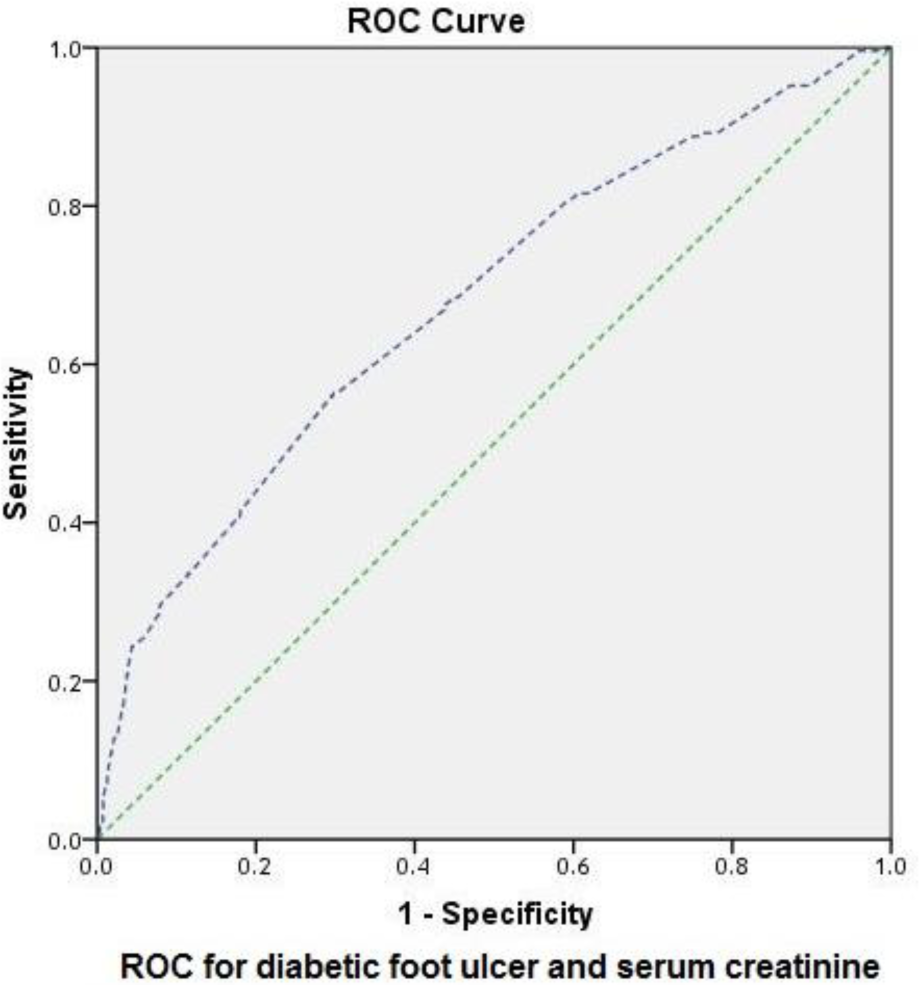

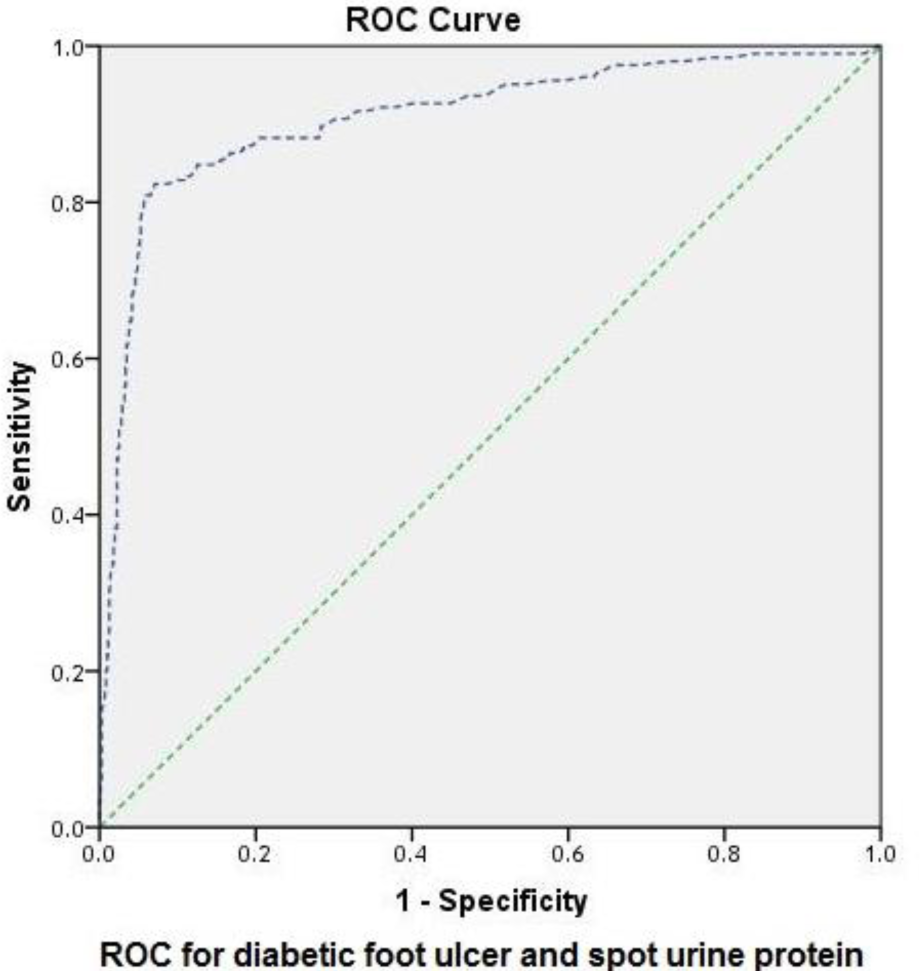

## DISCUSSION

Diabetic renal failure and nephropathy has been demonstrated to be associated with diabetic septic foot ulceration and amputation [56 57]. Additionally, diabetic patient on dialysis are also at risk of development of foot ulceration [58-63]. Moreover, uraemia and renal failure have been associated and are the risk factors for non-healing neuroischaemic foot lesions and amputations. Ureamia has a direct negative effect on ulcer healing as compared to nonuraemic patients [64-66]. Hence, in other words, DKD/CKD has strong association and is a risk for the development of DFU, chronic non-healing ulcers and amputations, vice versa [67]. It is well known that diabetes effects the kidney chronically and leads to decrease in the kidney function or glomerular filtration rate (GFR); and if untreated at earlier stages, may lead to ESRD [68-73]. Hence. this study investigated association of DFU with renal failure or DKD/CKD. Out of 10,680 patients, 12% presented with DFU. 43% was hypertensive. 39% demonstrated nephropathy, while 15% was diagnosed as DKD/CKD.

According to Table-3, we observed that levels of HbA1c, serum creatinine, systolic and diastolic BP, microalbumin in urine, spot urine protein and PCR were higher among the patients with DFU, with significant p-values. This statistical analysis suggests that elevated HbA1c or poor glycemic control contribute to the development of DFU and impairs wound healing. Furthermore, elevated BP significantly effects renal physiology with excretion of increased levels of microalbumin and proteins into the urine and development of nephropathy. All these pathophysiologic conditions and DFU are inter-related. Hence, patients with DFU has demonstrated elevated serum creatinine and renal impairment (DKD/CKD). Additionally, as demonstrated by Table-4, ABI values were lower among the patients demonstrating nephropathy and DKD/CKD (serum creatinine > 1.5 mg/dl), with significant p-values; this suggests that renal involvement in diabetic metabolic state is significantly associated with lower blood supply to the feet. This brings attention of clinical researchers to investigate this cause effect relationship at multi-center level [74,75]. Moreover, this data was further supported by conducting χ^2^ analysis in Table-5, which has demonstrated strong association of DFU with HTN, nephropathy and DKD/CKD (p-values < 0.0001 for all tested variables).

We also conducted statistical analysis in detail to find out cutoff points for HbA1c, serum creatinine and spot urine protein to detect the threshold levels of these variables which can significantly contribute to the development of DFU and can give indication to the physician that active intervention in required to prevent further complications. Hence, according to table-6, HbA1c values of 7.8% (g/dl) (with 60% sensitivity and 52% specificity), serum creatinine of 1.2 mg/dl (with 75% sensitivity and 48% specificity), and spot urine protein of 35 mg/dl (with 88% sensitivity and 90% specificity) was associated with development of DFU. Although microalbumin was studied in previous studies more extensively and was demonstrated to be a biomarker for CVD and incipient nephropathy.

**Table-6.** Results of ROC with AUC, 95% CI, p-values and triglyceride cutoff points

| **Test variables** | **Area**<br>**under the**<br>**curve**<br>**(AUC)** | **Standard**<br>**error** | **95% CI** | **P-Value** | **Coordinate blood pressure**<br>**cutoff points for the**<br>**development of diabetic**<br>**retinopathy** |
| --- | --- | --- | --- | --- | --- |
| Diabetic foot<br>ulcer and<br>HbA1c | 0.58 | 0.26 | 0.521 to 0.624 | < 0.0006 | 7.8 mmHg<br>(60% sensitivity and 52%<br>specificity) |
| Diabetic foot<br>ulcer and<br>creatinine | 0.68 | 0.19 | 0.640 to 0.715 | < 0.0001 | 1.2 mg/dl (75% sensitivity<br>and 48% specificity) |
| Diabetic foot<br>ulcer and spot<br>urine protein | 0.585 | 0.016 | 0.555 to 0.616 | < 0.0001 | 35 mg/dl<br>(88% sensitivity and 90%<br>specificity) |

However, our current study for the first time has demonstrated that spot urine protein excretion from the kidney is also as strong risk factor and biomarker for the development of nephropathy and DFU. Glycemic control should be optimal and HbA1c should be near the targets (7% to 7.5%) as current data has indicated that DFU was associated with elevated HbA1c (7.8%). Better glycemic control improves wound healing and prevents diabetic complications.

Finally, it can be concluded that diabetic patients should be assessed and screened at early stages in tertiary care diabetes centers for the detection of HTN, nephropathy, neuropathy or diabetic foot screening, dyslipidemia, and retinopathy as well to prevent complications and reduce health cost. Diabetes guidelines should be used to manage diabetes and its complications, including diabetic foot ulcers [76-98].

We have investigated for the first time risk factors such as elevated HbA1c, elevated BP or HTN, microalbuminuria, spot urine protein, for the development of DFU in the presence of nephropathy and DKD. We have also investigated association of low ABI with nephropathy and DKD. Our data analysis was in consistent with past studies. Further studies at multicenter level are required to confirm the results of the current study.

## CONCLUSION AND RECOMMENDATIONS

Our data has prompted and recommended diabetologists, endocrinologists, and physicians to use routine assessment and screening for diabetes complication detection. These include HTN, dyslipidemia, nephropathy, DKD, routine assessment of the feet and peripheral circulation at regular intervals and to focus especially on increasing serum creatinine, proteinuria and renal failure (DKD/CKD) among the patients with diabetic foot ulcer or DFI.

## Data Availability

From Diabetology Clinic

## REFERENCES

1. King H, Aubert RA, Herman WH. Global burden of diabetes, 1995-2025. Prevalence, numerical estimates, and projections. Diabetes Care 1998; 21:1414.

2. Reiber GE. Epidemiology of foot ulcers and amputations in the diabetic foot. In: Bowker JH, Pfeifer MA, (Eds) the Diabetic Foot, St. Louis: Mosby; 2001. p. 13–32.

3. Reiber GE, Boyko EJ, Smith DG. Lower extremity foot ulcers and amputations in diabetes. In: Harris MI, Cowie C, Stern MP, (Eds). Diabetes in America, 2nd ed, USA: NIH Publication; No. 95-1468; 1995. P. 409–27.

4. Prompers L, Huijberts M, Apelqvist J, Jude E, Piaggesi A, Bakker K, et al. High prevalence of ischaemia, infection and serious comorbidity in patients with diabetic foot disease in Europe: baseline results from the EURODIALE study. Diabetologia 2007; 50 : 18–25.

5. Boulton AJM, Vileikyte L, Ragnarson - Tennvall G, Apelqvist J. The global burden of diabetic foot disease. Lancet 2005; 366: 1719–1724.

6. Boulton AJM. The diabetic foot: from art to science: the 18^th^ Camillo Golgi lecture. Diabetologia 2004; 47: 1343 – 1353.

7. Connor H. Some historical aspects of diabetic foot disease. Diabet Metab Res Rev 2008; 24 (Suppl 1): 7–s13.

8. McKeown KC. The history of the diabetic foot. Diabet Med 1995; 12: 19 – 23.

9. ämaan A, Tajiyeva O, Müller N, Tschauner T, Hoyer H, Wolf G, et al Prevalence of the diabetic foot syndrome at the primary care level in Germany: a cross - sectional study. Diabet Med 2008; 25: 557 – 563.

10. Lavery LA, Armstrong DG, Wunderlich RP, Tredwell J, Boulton AJ. Diabetic foot syndrome: evaluating the prevalence and incidence of foot pathology in Mexican Americans and non-Hispanic whites from a diabetes disease management cohort. Diabetes Care 26:1435–1438, 2003.

11. Lobmann R, Schultz G, Lehnert H. Proteases and the diabetic foot syndrome: mechanisms and therapeutic implications. Diabetes Care 28:461–471, 2005.

12. Enderle MD, Coerper S, Schweizer HP, Kopp AE, Thelen MH, Meisner C, Pressler H, Becker HD, Claussen C, Haring HU, Luft D. Correlation of imaging techniques to histopathology in patients with diabetic foot syndrome and clinical suspicion of chronic osteomyelitis. The role of high-resolution ultrasound. Diabetes Care 22:294–299, 1999.

13. Diabetes Control and Complications Trial Research Group. Effect of intensive diabetes treatment on the development and progression of long-term complications in adolescents with insulin-dependent diabetes mellitus: Diabetes Control and Complications Trial. Diabetes Control and Complications Trial Research Group. J Pediatr. 1994;125:177–88.

14. Retnakaran R, Cull CA, Thorne KI, Adler AI, Holman RR; UKPDS Study Group. Risk factors for renal dysfunction in type 2 diabetes: U.K. Prospective Diabetes Study 74. Diabetes 2006;55:1832–1839

15. Fioretto P, Dodson PM, Ziegler D, Rosenson RS. Residual microvascular risk in diabetes: unmet needs and future directions. Nat Rev Endocrinol 2010;6:19–25

16. National Kidney Foundation: KDOQI clinical practice guidelines and clinical practice recommendations for diabetes and chronic kidney disease. Am J Kidney Dis 2007; 49: (Suppl1) S180.

17. Molitch ME, DeFronzo RA, Franz MJ, Keane WF, Mogensen CE, Parving HH. American Diabetes Association. Diabetic nephropathy. Diabetes Care 2003;26(Suppl. 1):S94–S98

18. Mogensen CE. Definition of diabetic renal disease in insulin dependent diabetes mellitus based on renal function tests: The Kidney and Hypertension in Diabetes Mellitus. Kluwer Academic Publishers, Boston, USA, 1994:1–14.

19. American Diabetes Association. Cardiovascular disease and risk management. Diabetes Care 2016 40: S75–S87.

20. Cao C, Wan X, Chen Y, Wu W. Metabolic factors and micro inflammatory state promote kidney injury in type 2 diabetes mellitus patients. Ren Fail. 2009;31:470–474.

21. Cusick M, Chew EY, Hoogwerf B****, Agr?n E, Wu L, Lindley A, Ferris FL 3rd; Early Treatment Diabetic Retinopathy Study Research Group. Risk factors for renal replacement therapy in the Early Treatment Diabetic Retinopathy Study (ETDRS), Early Treatment Diabetic Retinopathy Study Report No. 26. Kidney Int. 2004;66:1173–1179.

22. Muntner P, Coresh J, Smith JC, Eckfeldt J, Klag MJ. Plasma lipids and risk of developing renal dysfunction: the Atherosclerosis Risk in Communities study. Kidney Int. 2000;58:293–301.

23. Ritz E, Rychlik I, Locatelli F, Halimi S: End stage renal failure in type 2 diabetes: a medical catastrophe of worldwide dimensions. Am J Kidney Dis 34:795–808, 1999.

24. Schena FP. Epidemiology of end stage renal disease: international comparisons of renal replacement therapy. Kidney Int Suppl 2000;57:S39–S45

25. Papamichael CM, Lekakis JP, Stamatelopoulos KS, Papaioannou TG, Alevizaki MK, Cimponeriu AT, Kanakakis JE, Papapanagiotou A, Kalofoutis AT, Stamatelopoulos SF. Ankle-brachial index as a predictor of the extent of coronary atherosclerosis and cardiovascular events in patients with coronary artery disease. The American journal of cardiology. 2000 Sep 15;86(6):615–8.

26. Heald CL, Fowkes FG, Murray GD, Price JF, Ankle Brachial Index Collaboration. Risk of mortality and cardiovascular disease associated with the ankle-brachial index: systematic review. Atherosclerosis. 2006 Nov 1;189(1):61–9.

27. Zheng ZJ, Sharrett AR, Chambless LE, Rosamond WD, Nieto FJ, Sheps DS, Dobs A, Evans GW, Heiss G. Associations of ankle-brachial index with clinical coronary heart disease, stroke and preclinical carotid and popliteal atherosclerosis:: the Atherosclerosis Risk in Communities (ARIC) Study. Atherosclerosis. 1997 May 1;131(1):115–25.

28. McDermott MM, Liu K, Greenland P, Guralnik JM, Criqui MH, Chan C, Pearce WH, Schneider JR, Ferrucci L, Celic L, Taylor LM. Functional decline in peripheral arterial disease: associations with the ankle brachial index and leg symptoms. Jama. 2004 Jul 28;292(4):453–61.

29. McDermott MM, Criqui MH, Liu K, Guralnik JM, Greenland P, Martin GJ, Pearce W. Lower ankle/brachial index, as calculated by averaging the dorsalis pedis and posterior tibial arterial pressures, and association with leg functioning in peripheral arterial disease. Journal of Vascular Surgery. 2000 Dec 1;32(6):1164–71.

30. Ix JH, Katz R, De Boer IH, Kestenbaum BR, Allison MA, Siscovick DS, Newman AB, Sarnak MJ, Shlipak MG, Criqui MH. Association of chronic kidney disease with the spectrum of ankle brachial index: the CHS (Cardiovascular Health Study). Journal of the American College of Cardiology. 2009 Sep 22;54(13):1176–84.

31. Kitahara T, Ono K, Tsuchida A, Kawai H, Shinohara M, Ishii Y, Koyanagi H, Noguchi T, Matsumoto T, Sekihara T, Watanabe Y. Impact of brachial-ankle pulse wave velocity and ankle-brachial blood pressure index on mortality in hemodialysis patients. American Journal of Kidney Diseases. 2005 Oct 1;46(4):688–96.

32. Caramori M, Fioretto P, Mauer M. The need for early predictors of diabetic nephropathy risk: Is albumin excretion rate sufficient?. Diabetes 2000; 49: 1399–408.

33. Nosadini R, Velussi M, Brocco E, Bruseghin M, Abaterusso C, Saller A, et al. Course of renal function in type 2 diabetic patients with abnormalities of albumin excretion rate. Diabetes 2000; 49: 476–84.

34. Tabaei B, Al-Kassab A, Ilag L, Zawacki C, Herman W. Does microalbuminuria predict diabetic nephropathy?. Diabetes Care 2001; 24: 1560–6.

35. Fioretto P, Mauer M, Brocco E, Velussi M, Frigato F, Muollo B, et al. Patterns of renal injury in NIDDM patients with microalbuminuria. Diabetologia 1996; 39: 1569–76.

36. Kramer H, Nguyen Q, Curhan G, Hsu C. Renal insufficiency in the absence of albuminuria and retinopathy among adults with type 2 diabetes mellitus. JAMA 2003; 289: 3273–7.

37. Remuzzi G, Schieppati A, Ruggenenti P. Clinical practice: Nephropathy in patients with type 2 diabetes. N Engl J Med 2002; 346:1145–51.

38. Chavers BM, Bilous RW, Ellis EN, Steffes MW, Mauer SM. Glomerular lesions and urinary albumin excretion in type 1 diabetes without overt proteinuria. N EnglJ Med 1989; 320: 966 –70.

39. Aziz KMA. Correlation of urine biomarkers: microalbuminuria and spot urine protein among diabetic patients. application of spot urine protein in diabetic kidney disease, nephropathy, proteinuria estimation, diagnosing and monitoring. Recent patents on endocrine, metabolic & immune drug discovery. 2015 Aug 1;9(2):121–33.

40. Rodby R, Rohde R, Sharon Z, Pohl M, Bain R, Lewis E. The urine protein to creatinine ratio as a predictor of 24-hour urine protein excretion in type 1 diabetic patients with nephropathy. The Collaborative Study Group. Am J Kidney Dis 1995; 26: 904–9.

41. Eddy A, McCulloch L, Liu E, Adams J. A relationship between proteinuria and acute tubulo-interstitial disease in rats with experimental nephrotic syndrome. Am J Pathol 1991;138: 1111–23.

42. Côté A, Brown M, Lam E, von Dadelszen P, Firoz T, Liston R, et al. Diagnostic accuracy of urinary spot protein:creatinine ratio for proteinuria in hypertensive pregnant women: Systematic review. BMJ 2008; 336:1003–6.

43. Lemann J, Doumas B. Proteinuria in health and disease assessed by measuring the urinary protein/creatinine ratio. Clin Chem 1987; 33:297–9.

44. Ginsberg J, Chang B, Matarese R, Garella S. Use of single voided urine samples to estimate quantitative proteinuria. N Engl J Med 1983; 309: 1543–6.

45. Ruggenenti P, Gaspari F, Perna A, Remuzzi G. Cross sectional longitudinal study of spot morning urine protein:creatinine ratio, 24 hour urine protein excretion rate, glomerular filtration rate, and end stage renal failure in chronic renal disease in patients without diabetes. BMJ 1998; 316: 504–9.

46. Abitbol C, Zilleruelo G, Freundlich M, Strauss J. Quantitation of proteinuria with urinary protein/creatinine ratios and random testing with dipsticks in nephrotic children. J Pediatr 1990; 116: 243–7.

47. Morgenstern B, Butani L, Wollan P, Wilson D, Larson T. Validity of protein-osmolality versus protein-creatinine ratios in the estimation of quantitative proteinuria from random samples of urine in children. Am J Kidney Dis 2003; 41: 760–6.

48. Kim H, Cheon H, Choe J, Yoo K, Hong Y, Lee J, et al. Quantification of proteinuria in children using the urinary protein-osmolality ratio. Pediatr Nephrol. 2001;16:73–6

49. Houser M. Assessment of proteinuria using random urine samples. J Pediatr 1984;104: 845–8.

50. Margolis DJ, Hofstad O, Feldman HI. The association between renal failure and foot ulcer or lower extremity amputation in those patients with diabetes. Diabetes care. 2008 Apr 3.

51. Mitchell RJ. Improved method for specific determination of creatinine in serum and urine. Clin Chem 1973; 19: 408–10.

52. Slot C. Plasma creatinine determination. A new and specific Jaffe reaction method. Scand J Clin Lab Invest 1965; 17: 381–7.

53. Bishop Michael L. Clinical Chemistry: Principles and Correlations 2nd ed. Philadelphia: Lippincott JB, Company 1992; 441.

54. Fujita Y, Mori I, Kitano S. Color reaction between pyrogallol red molybdenum (VI) complex and protein. Bunseki Kagaku 1983; 32:379–86.

55. NATCOM Hospital Information System (NATCOM HIS), National Computer System Co, Ltd. http://natcom.com.sa/healthcare and http://natcom.com.sa/clients (Accessed on: April 17, 2018).

56. Fernando DJ, Hutchison A, Veves A, Gokal R, Boulton AJ. Risk factors for non-ischaemic foot ulceration in diabetic nephropathy. Diabetic medicine. 1991 Apr;8(3):223–5.

57. Guerrero-Romero F, Rodr guez-Morán M. Relationship of microalbuminuria with the diabetic foot ulcers in type II diabetes. Journal of Diabetes and its Complications. 1998 Jul 1;12(4):193–6.

58. Ndip A, Rutter MK, Vileikyte L, Vardhan A, Asari A, Jameel M, Tahir HA, Lavery LA, Boulton AJ. Dialysis treatment is an independent risk factor for foot ulceration in patients with diabetes and stage 4 or 5 chronic kidney disease. Diabetes care. 2010 May 19

59. Dòria M, Rosado V, Pacheco LR, Hernández M, Betriu Á, Valls J, Franch-Nadal J, Fernández E, Mauricio D. Prevalence of diabetic foot disease in patients with diabetes mellitus under renal replacement therapy in Lleida, Spain. BioMed research international. 2016;2016.

60. Morbach S, Quante C, Ochs HR, Gaschler F, Pallast JM, Knevels U: Increased risk of lower-extremity amputation among Caucasian diabetic patients on dialysis. Diabetes Care 24:1689–1690, 2001

61. O’Hare AM, Bacchetti P, Segal M, Hsu CY, Johansen KL, Dialysis Morbidity and Mortality Study Waves: Factors associated with future amputation among patients undergoing hemodialysis: results from the Dialysis Morbidity and Mortality Study Waves 3 and 4. Am J Kidney Dis 41:162–170, 2003

62. O’Hare AM, Glidden DV, Fox CS, Hsu CY: High prevalence of peripheral arterial disease in persons with renal insufficiency: results from National Health and Nutrition Examination Survey 1999–2000. Circulation 109:320–323, 2004

63. O’Hare AM, Vittinghoff E, Hsia J, Shlipak MG: Renal insufficiency and the risk of lower extremity peripheral arterial disease: results from Heart and Estrogen/Progestin Replacement Study. J Am Soc Nephrol 15:1046–1051, 2004

64. Prompers L, Schaper N, Apelqvist J, Edmonds M, Jude E, Mauricio D, Uccioli L, Urbancic V, Bakker K, Holstein P, Jirkovska A. Prediction of outcome in individuals with diabetic foot ulcers: focus on the differences between individuals with and without peripheral arterial disease. The EURODIALE Study. Diabetologia. 2008 May 1;51(5):747–55.

65. Gershater MA, Löndahl M, Nyberg P, Larsson J, Thörne J, Eneroth M, Apelqvist J. Complexity of factors related to outcome of neuropathic and neuroischaemic/ischaemic diabetic foot ulcers: a cohort study. Diabetologia. 2009 Mar 1;52(3):398–407.

66. Dinh TL, Veves A. A review of the mechanisms implicated in the pathogenesis of the diabetic foot. The international journal of lower extremity wounds. 2005 Sep;4(3):154–9.

67. Wolf G, Müller N, Busch M, Eidner G, Kloos C, Hunger-Battefeld W, Müller UA. Diabetic foot syndrome and renal function in type 1 and 2 diabetes mellitus show close association. Nephrology Dialysis Transplantation. 2009 Jan 7;24(6):1896–901.

68. Mogensen CE. Glomerular filtration rate and renal plasma flow in short-term and long-term juvenile diabetes mellitus. Scandinavian journal of clinical and laboratory investigation. 1971 Jan 1;28(1):91–100.

69. O’Bryan GT, Hostetter TH. The renal hemodynamic basis of diabetic nephropathy. InSeminars in nephrology 1997 Mar (Vol. 17, No. 2, pp. 93–100).

70. Bethesda MD. US Renal Data System. USRDS 2005 Annual Data Report. National Institutes of Health, National Institute of Diabetes and Digestive and Kidney Diseases. 2005.

71. Bohlender JM, Franke S, Stein G, Wolf G. Advanced glycation end products and the kidney. American Journal of Physiology-Renal Physiology. 2005 Oct;289(4):F645–59.

72. Rüster C, Bondeva T, Franke S, Förster M, Wolf G. Advanced glycation end-products induce cell cycle arrest and hypertrophy in podocytes. Nephrology Dialysis Transplantation. 2008 Mar 14;23(7):2179–91.

73. Thomson SC, Vallon V, Blantz RC. Kidney function in early diabetes: the tubular hypothesis of glomerular filtration. American Journal of Physiology-Renal Physiology. 2004 Jan;286(1):F8–15.

74. Garimella PS, Hart PD, O’Hare A, DeLoach S, Herzog CA, Hirsch AT. Peripheral artery disease and CKD: a focus on peripheral artery disease as a critical component of CKD care. American Journal of Kidney Diseases. 2012 Oct 1;60(4):641–54.

75. Lepäntalo M, Fiengo L, Biancari F. Peripheral arterial disease in diabetic patients with renal insufficiency: a review. Diabetes/metabolism research and reviews. 2012 Feb;28:40–5.

76. Aziz KMA. Management of type-1 and type-2 diabetes by insulin injections in diabetology clinics-a scientific research review. Recent patents on endocrine, metabolic & immune drug discovery. 2012 May 1;6(2):148–70.

77. American Diabetes Association. 6. Glycemic targets: standards of medical care in diabetes—2018. Diabetes Care. 2018 Jan 1;41(Supplement 1):S55–64.

78. Aziz KMA. Unique glycemic and cardio-renal protective effects of metformin therapy among type-2 diabetic patients: a lesson from a five-year cross-sectional observational study of 1590 patients. Research. 2014 Jun 12.

79. Muller UA, Femerling M, Reinauer KM, Risse A. Intensified treatment and education of type 1 diabetes as clinical routine. Diabetes Care. 1999 Mar 1;22:B29.

80. Aziz KMA. Association of microalbuminuria with ischemic heart disease, dyslipidemia and obesity among diabetic patients: Experience from 5 year follow up study of 1415 patients. Bioenergetics. 2014;3:118.

81. Aziz KMA, Al-Qahtani MA. Association between Non-HDL and HDL Cholesterol with microalbuminuria in patients with Diabetes. Journal of Diabetology. 2013 Feb;1(4).

82. Stone NJ, Robinson JG, Lichtenstein AH, Merz CN, Blum CB, Eckel RH, Goldberg AC, Gordon D, Levy D, Lloyd-Jones DM, McBride P. 2013 ACC/AHA guideline on the treatment of blood cholesterol to reduce atherosclerotic cardiovascular risk in adults: a report of the American College of Cardiology/American Heart Association Task Force on Practice Guidelines. Journal of the American College of Cardiology. 2014 Jul 1;63(25 Part B):2889–934.

83. Aziz KMA. Association between Hypothyroidism, Body Mass Index, Systolic Blood Pressure and Proteinuria in Diabetic Patients: Does treated Hypothyroid with Thyroxine Replacement Therapy prevent Nephropathy/ Chronic Renal Disease? Current Diabetes Reviews, 12(3): 297–306.

84. Aziz KMA. (2018) Association of Diabetic Retinopathy and Maculopathy with Elevated HbA1c, Blood Pressure, Serum Creatinine, Microalbuminuria, Spot Urine Protein, Nephropathy and Diabetic Kidney Disease. An Experience from Data Analysis of 10,580 Diabetic Patients.J Endocrinol Diab. 5(1): 1–11.

85. McCabe CJ, Stevenson RC, Dolan AM. Evaluation of a diabetic foot screening and protection programme. Diabetic Medicine. 1998 Jan;15(1):80–4.

86. Aziz KMA. The Diabetic Foot Syndrome an Ignored and Potential Problem in Medical Practice. International Journal of Diabetology & Vascular Disease Research. 2013:1–3.

87. Treece KA, Macfarlane RM, Pound N, Game FL, Jeffcoate WJ. Validation of a system of foot ulcer classification in diabetes mellitus. Diabetic medicine. 2004 Sep;21(9):987–91.

88. Bakker K, Apelqvist J, Lipsky BA, Van Netten JJ, Schaper NC, International Working Group on the Diabetic Foot (IWGDF). The 2015 IWGDF guidance documents on prevention and management of foot problems in diabetes: development of an evidence-based global consensus. Diabetes/metabolism research and reviews. 2016 Jan;32:2–6.

89. Bus SA, Van Netten JJ, Lavery LA, Monteiro-Soares M, Rasmussen A, Jubiz Y, Price PE, International Working Group on the Diabetic Foot (IWGDF). IWGDF guidance on the prevention of foot ulcers in at-risk patients with diabetes. Diabetes/metabolism research and reviews. 2016 Jan;32:16–24.

90. Aziz KMA. Association of Hypothyroidism with High Non-HDL Cholesterol and Ankle Brachial Pressure Index in Patients with Diabetes: 10-Year Results from a 5780 Patient Cohort. A Need for Intervention. Annals Thyroid Res. 2016; 2(2): 53–57.

91. Wagner Jr FW. The dysvascular foot: a system for diagnosis and treatment. Foot & ankle. 1981 Sep;2(2):64–122.

92. Armstrong DG, Lavery LA, Harkless LB. Validation of a diabetic wound classification system: the contribution of depth, infection, and ischemia to risk of amputation. Diabetes care. 1998 May 1;21(5):855–9.

93. Lavery LA, Armstrong DG, Vela SA, Quebedeaux TL, Fleischli JG. Practical criteria for screening patients at high risk for diabetic foot ulceration. Archives of internal medicine. 1998 Jan 26;158(2):157–62.

94. Aziz KMA. Association of high serum triglycerides and triglycerides/HDL ratio with raised HbA1c, creatinine, microalbuminuria and development of diabetic kidney disease and diabetic renal failure. Mathematical and statistical regression models of 10,370 diabetic patients. Clinical Nephrology and Research. 2017 Dec 22;1(1).

95. Wolf G, Ritz E. Diabetic nephropathy in type 2 diabetes prevention and patient management. Journal of the American society of nephrology. 2003 May 1;14(5):1396–405.

96. Aziz KMA. Targeting LDL Dyslipidemia for Controlling Progression of Nephropathy in Diabetic Population: A Cross Sectional Analytical Study. Journal of Dow University of Health Sciences. 2013 Jun 10;6(1).

97. Vanholder R. Chronic kidney disease in adults—UK guidelines for identification, management and referral. Nephrology Dialysis Transplantation. 2006 Jul 1;21(7):1776–7.

98. Aziz KMA. Association of Serum Lipids with High Blood Pressure and Hypertension among Diabetic Patients. Mathematical Regression Models to Predict Blood Pressure from Lipids. An Experience from 12-year Follow Up of more than 9000 Patients’ Cohort. Gen Med (Los Angeles) 5: 297. doi:10.4172/2327-5146.1000297

